# Fatal COVID-19 outcomes are associated with an antibody response targeting epitopes shared with endemic coronaviruses

**DOI:** 10.1101/2021.05.04.21256571

**Authors:** Anna L McNaughton, Robert S Paton, Matthew Edmans, Jonathan Youngs, Judith Wellens, Prabhjeet Phalora, Alex Fyfe, Sandra Belij-Rammerstorfer, Jai S Bolton, Jonathan Ball, George Carnell, Wanwisa Dejnirattisai, Christina Dold, David W Eyre, Philip Hopkins, Alison Howarth, Kreepa Kooblall, Hannah Klim, Susannah Leaver, Lian Lee, César López-Camacho, Sheila F Lumley, Derek Macallan, Alexander J Mentzer, Nicholas Provine, Jeremy Ratcliff, Jose Slon-Compos, Donal Skelly, Lucas Stolle, Piyada Supasa, Nigel Temperton, Chris Walker, Beibei Wang, Duncan Wyncoll, OPTIC consortium, SNBTS consortium, Peter Simmonds, Teresa Lambe, Kenneth Baillie, Malcolm G Semple, Peter IM Openshaw, ISARIC4C Investigators, Uri Obolski, Marc Turner, Miles Carroll, Juthathip Mongkolsapaya, Gavin Screaton, Stephen H Kennedy, Lisa Jarvis, Eleanor Barnes, Susanna Dunachie, José Lourenço, Philippa C Matthews, Tihana Bicanic, Paul Klenerman, Sunetra Gupta, Craig P Thompson

**Affiliations:** Peter Medawar Building for Pathogen Research, South Parks Road, Oxford, UK; Nuffield Department of Medicine, University of Oxford, Oxford, UK; Department of Zoology, University of Oxford, Oxford, United Kingdom; Institute of Infection & Immunity, St. George’s University of London, London, UK; Translational Gastro-intestinal Unit, Nuffield Department of Medicine, John Radcliffe Hospital, Oxford; Translational Research for Gastrointestinal Diseases, University hospitals Leuven, Herestraat, Leuven, Belgium; The Jenner Institute Laboratories, University of Oxford, Oxford, UK; Intensive Care Medicine, St. George’s University Hospital NHS Trust, London UK; Department of Veterinary Medicine, University of Cambridge, Cambridge,, UK; Wellcome Centre for Human Genetics, Nuffield Department of Medicine, University of Oxford, Oxford, UK; Oxford Vaccine Group, Oxford Vaccine Group, Department of Paediatrics, University of Oxford, UK; Nuffield Department of Population Health, University of Oxford, Oxford, UK; Centre for Human & Applied Physiological Sciences, School of Basic & Medical Biosciences, Faculty of Life Sciences, & Medicine, King’s College, London, UK; Department of Microbiology/Infectious Diseases, Oxford University Hospitals NHS Foundation Trust, John Radcliffe Hospital, Oxford, UK; Oxford Centre for Diabetes, Endocrinology and Metabolism, Churchill Hospital, University of Oxford, Oxford, UK; Future of Humanity Institute, Department of Philosophy, University of Oxford, UK; Nuffield Department of Clinical Neurosciences, University of Oxford; Oxford University Hospitals NHS Foundation Trust; Department of Biochemistry, University of Oxford, Oxford, UK; Viral Pseudotype Unit, Medway School of Pharmacy, University of Kent, Chatham, UK; Meso-Scale Diagnostics, Maryland, USA; Intensive Care Medicine, Guys and St Thomas’ Hospital NHS Foundation Trust, London, UK; Health Protection Unit in Emerging and Zoonotic Infection, Faculty of Health and Life Sciences, University of Liverpool, Liverpool, UK; NIHR Health Protection Research Unit in Emerging and Zoonotic Infections, Faculty of Health and Life Sciences, University of Liverpool, Liverpool, UK; National Heart and Lung Institute, Imperial College, London, UK; School of Public Health, Faculty of Medicine, Tel-Aviv University, Tel-Aviv, Israel; Porter School of Environmental and Earth Sciences, Faculty of Exact Sciences, Tel- Aviv University, Tel-Aviv, Israel; National Microbiology Reference Unit, Scottish National Blood Transfusion Service, Edinburgh, UK; National Infection Service, Public Health England (PHE), Porton Down, Salisbury, UK; Siriraj Center of Research Excellence in Dengue & Emerging Pathogens, Dean Office for Research, Faculty of Medicine Siriraj Hospital, Mahidol University, Thailand; Chinese Academy of Medical Science (CAMS) Oxford Institute (COI), University of Oxford, Oxford, UK; Nuffield Department of Women’s & Reproductive Health, University of Oxford, John Radcliffe Hospital, Oxford, United Kingdom; Centre for Tropical Medicine and Global Health, Nuffield Department of Medicine, University of Oxford, UK

**Author notes:** Corresponding author: Craig Thompson. These authors contributed equally (Anna L McNaughton, Robert S Paton).

## Abstract

It is unclear whether prior endemic coronavirus infections affect COVID-19 severity. Here, we show that in cases of fatal COVID-19, antibody responses to the SARS-COV-2 spike are directed against epitopes shared with endemic beta-coronaviruses in the S2 subunit of the SARS-CoV-2 spike protein. This immune response is associated with the compromised production of a *de novo* SARS-CoV-2 spike response among individuals with fatal COVID-19 outcomes.

## Introduction

Four human coronaviruses (HCoV) are currently considered endemic worldwide. These include two beta-coronaviruses, HCoV-OC43 and HCoV-HKU1, as well as two alpha-coronaviruses, HCoV-229E and HCoV-NL63. Infection by these viruses in the majority of people causes a mild respiratory illness (1). Over the past two decades, two further beta-coronaviruses have also emerged, SARS-CoV-1 and MERS-CoV. Whilst both viruses have been more pathogenic than endemic coronaviruses, their transmission and subsequent spread has remained limited (2). In 2019 a fifth beta-coronavirus, SARS-CoV-2, emerged which has led to a pandemic with over 100 million cases and upwards of 3 million deaths confirmed to date (3). Several studies have shown that prior infection with other HCoVs induces both cross-reactive antibody and T-cell cross-reactive responses to SARS-CoV-2 (4–7). However, the influence of prior HCoV exposure on COVID-19 severity and clinical outcome remains unclear (8).

The spike protein of SARS-CoV-2, which is the primary vaccine target, consists of the S1 and S2 subunits (9,10). The S1 subunit contains a more variable receptor-binding domain (RBD), which mediates viral entry during the infection process via interaction with the angiotensin-converting enzyme 2 (ACE2) receptor. Antibodies targeting the RBD can be neutralising and have been shown to correlate with protection (11). HCoV induced antibody responses do not appear to cross-react with the SARS-CoV-2 RBD (9,12), or at least do so infrequently (10). In contrast, it has previously been reported by several studies that antibodies induced by prior HCoV infections cross-react with the SARS-CoV-2 S2 subunit (7,8,9,10).

Cross-reactive T-cell responses have also been reported to be present in many individuals (5,7,16,17), which may have been induced by HCoV infection. These studies found CD4+ and CD8+ T cells reactive to SARS-CoV-2 spike peptide pools in blood samples from individuals unexposed to SARS-CoV-2 (5,7,16). This indicates that prior exposure to HCoVs could confer a cross-reactive T-cell response in a subset of the population and could potentially affect COVID-19 severity (18).

In this study, to determine if prior infection by HCoVs is associated with differences in COVID-19 severity, we retrospectively tested samples from individuals who previously had qRT-PCR-confirmed asymptomatic infection, as well as patients admitted to ICU with severe COVID-19, half of whom died.

We also analysed two large cohorts with SARS-CoV-2 neutralising antibodies obtained from UK seroprevalence studies: one containing sera from blood donors and the other sera collected from pregnant women sampled <14 weeks’ gestation (19,20). These two cohorts did not have a precise clinical definition of SARS-CoV-2 infection severity. As a third control cohort, we included SARS-CoV-2 seronegative individuals from the same blood donor seroprevalence study (20). Further details of the cohorts can be found in Table 1 and Figures S3 to S5.

**Table 1.** Features of sample cohorts analysed.

| Cohort | N | Identification | Sampling stage | Clinical features | Sex (Female, n, %) | Age (Median, yrs, IQR) |
| --- | --- | --- | --- | --- | --- | --- |
| Scottish Blood Donors (background HCoV exposure samples) | 50 | Absence of anti-SARS-CoV-2 neutralising antibodies | During routine blood donation | No SARS-CoV-2 infection. Collected between March and May 2020. | 24, 48% | 44, 28 |
| Scottish blood donors (SARS-2 seropositive samples) | 109 | Detection of anti-SARS-CoV-2 neutralising antibodies (in-house SARS-CoV-2 spike pseudotype neutralisation assay) | During routine blood donation | Evidence of previous SARS-CoV-2 infection, disease course assumed mild-asymptomatic. | 50, 45.9% | 49, 25 |
| Antenatal samples (SARS-2 seropositive samples) | 56 | Detection of anti-SARS-CoV-2 neutralising antibodies (n=39)<br>ELISA positive for anti-SARS-2 antibodies (n=53) | During routine antenatal screening in the first trimester, generally at 8-12 weeks | Evidence of previous SARS-CoV-2 infection, disease course assumed mild-asymptomatic. | 56, 100% | NA |
| Asymptomatic samples | 22 | PCR-positive for SARS-CoV-2 | Sampled after confirmation of PCR-positive result | Confirmed SARS-CoV-2 infection, absence of clinical symptoms (from the ISARIC4C and PITCH studies). | NA | NA |
| Intensive Care Unit (ICU) samples | 42 | PCR-positive for SARS-CoV-2 | Sampled at admission to ICU | Confirmed SARS-CoV-2 infection, patients were admitted to ICU with severe COVID-19. A fatal outcome was recorded in 21/42 cases (from the AspiFlu study). | 9, 42.9% (fatal)<br>6, 28.6% (non-fatal) | 62, 15 (fatal)<br>50, 16 (non-fatal) |

By comparing antibody responses to the SARS-CoV-2 nucleocapsid and spike proteins, we show that individuals with fatal COVID-19 outcomes have an immune response directed against immunologically similar SARS-CoV-2 and beta-HCoV epitopes in the S2 subunits of the SARS-CoV-2 spike protein. This immune response is associated with the compromised generation of a *de novo* immune response to the SARS-CoV-2 spike in individuals with fatal COVID-19 outcomes. To our knowledge this is the first-time prior exposure to HCoVs has been associated with COVID-19 severity and mapped to a specific location on the spike protein.

## Results

### Individuals with fatal COVID-19 outcomes make lower responses to the SARS-CoV-2 spike protein but not the SARS-CoV-2 nucleocapsid

We used an MSD V-PLEX assay to quantify total antibody responses to the SARS-CoV-2 nucleocapsid (N), the RBD, N-terminal domains (NTD) of the spike, the SARS-CoV-2 full-length spike as well as the spike proteins of the four HCoVs and SARS-CoV-1. In-house indirect ELISAs were also developed for the SARS-CoV-2 RBD and full-length spike, in addition to the full-length alpha- and beta-HCoV spike proteins to confirm the results produced by the V-PLEX assay. Both the MSD assay and in-house ELISAs correlated well (Figure S1).

We found that the antibody titres to the SARS-CoV-2 spike and nucleocapsid generally correlated with increasing COVID-19 severity (Figure 1). However, individuals with fatal COVID-19 consistently exhibited lower titres to SARS-CoV-2 spike antigens; the SARS-CoV-2 RBD (p = 0.01), and NTD (p = 0.02) of the spike as well as the full length spike (p = 0.02) were higher in the non-fatal ICU cohort compared to the fatal ICU cohort (Figure 1a). In contrast, no difference in reactivity to the SARS-CoV-2 nucleocapsid protein was identified (p = 0.99).

**Figure 1:**
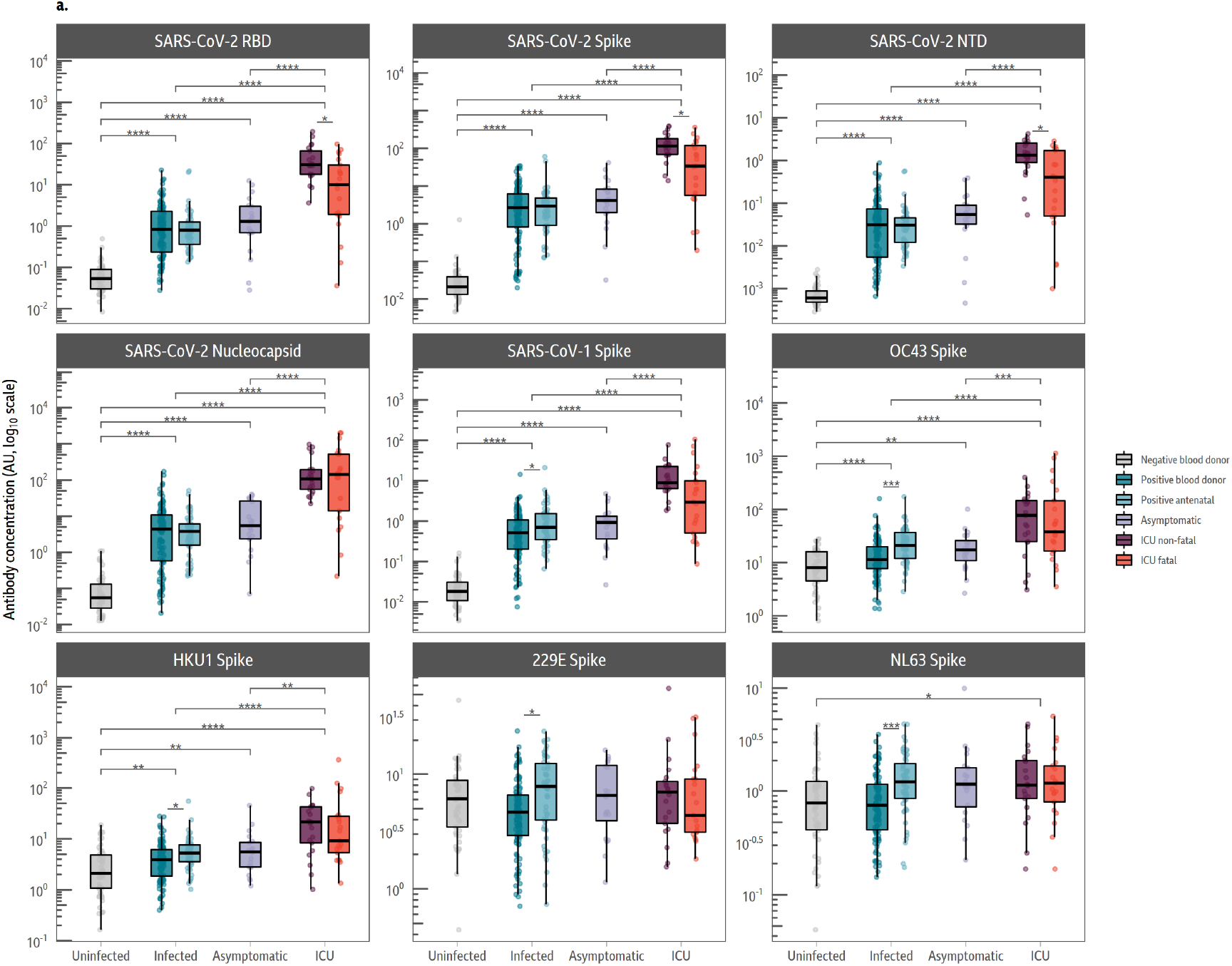
Individuals with fatal COVID-19 make lower responses to the SARS-CoV-2 spike protein, but not the SARS-CoV-2 nucleocapsid. **a. Boxplots comparing antibody concentrations for each antigen.** Sample groups (background uninfected, infected, asymptomatic and ICU) are given on the x-axis. Sub-groups are denoted by colour. The average response to all SARS-CoV-2 antigens were elevated in ICU patients, and no differences were observed between the infected and asymptomatic groups. Fatal ICU patients made a lower response to SARS-CoV-2 RBD (p = 0.01), spike (p = 0.02) and NTD (p = 0.02) than non-fatal ICU patients. The data in this figure were generated using the MSD V-PLEX assay.

To test responses to the S2 subunit of the SARs-CoV-2 spike protein we developed an S2 indirect ELISA. In contrast to the other SARS-CoV-2 spike antigens measured by both the V-PLEX assay and in-house ELISA, there was no difference in SARS-CoV-2 S2 ELISA responses the ICU fatal and non-fatal cohorts (Figure 2a: p = 0.99).

**Figure 2:**
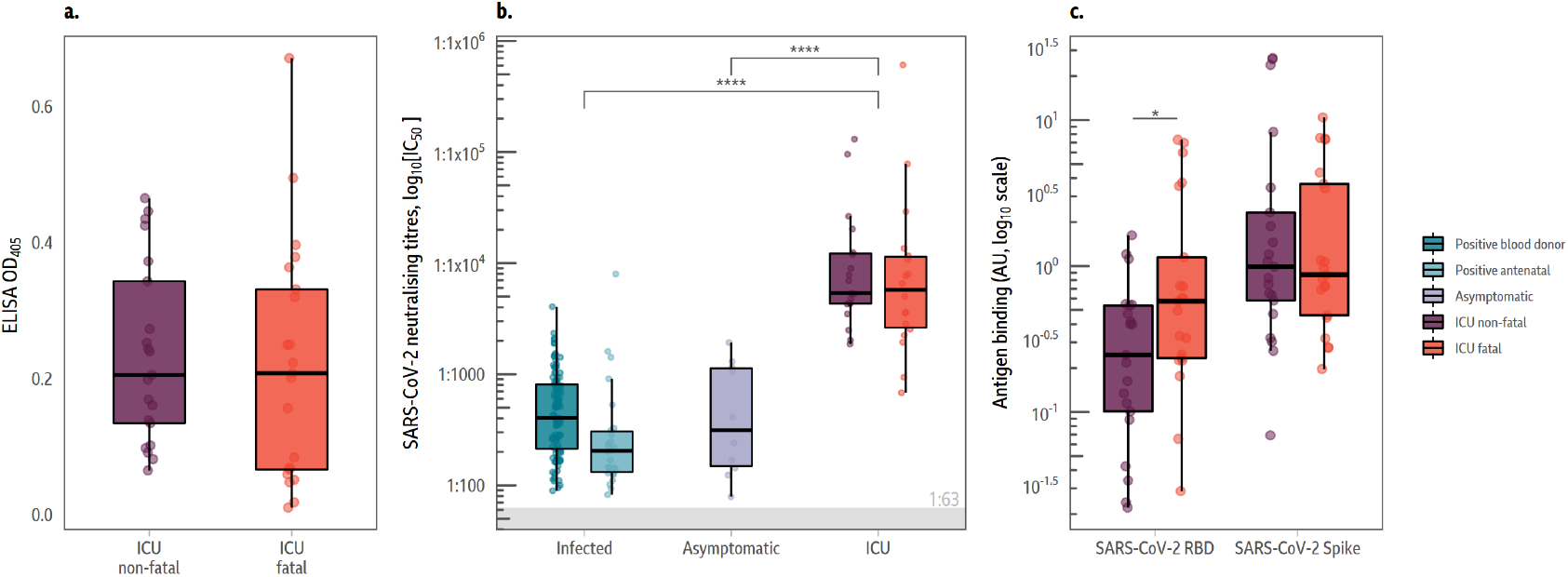
S2 ELISA responses and neutralising antibody responses cannot distinguish between severe non-fatal and fatal COVID-19. **a. Antibody responses directed against the S2 subunit of the SARS-CoV-2 spike protein.** There was no statistical difference between antibody responses directed at the S2 subunit of the SARS-CoV-2 spike in individuals with fatal and non-fatal COVID-19. Data generated by indirect ELISA. **b. Neutralising antibody levels correlate with disease severity but cannot distinguish between severe non-fatal and fatal COVID-19**. Samples were tested using a SARS-CoV-2 pseudotype microneutralisation assay. Neutralisation titres were higher in the ICU cohorts than the blood donor, antenatal and asymptomatic groups, but there was no significant difference based on clinical outcomes of ICU stay (p = 0.99). **c. ACE2 inhibition assay results**. Samples were also analysed with an R-PLEX ACE2 inhibition assay (MSD). The level of ACE2 binding inhibition was not statistically significant for the spike, but the fatal ICU cohort shows statistically lower ACE2-RBD binding inhibition in comparison to the non-fatal ICU cohort (p = 0.02).

Fatal COVID-19 outcomes were not associated with lower neutralising antibody titres as measured using a pseudotyped SARS-CoV-2 virus microneutralisation assay (Figure 2b, p = 0.96). An R-PLEX ACE2 competition assay using the full-length spike and RBD as antigens was run to confirm this result. The R-PLEX assay showed no significant difference in binding inhibition between ACE2 and the spike protein (p = 0.83). However, there was significantly lower inhibition of binding between the RBD and ACE2 in the R-PLEX for individuals with fatal outcomes (Figure 2c; p = 0.02).

### SARS-CoV-2 infection boosts responses to endemic beta-coronaviruses relative to disease severity

We compared responses to HCoV spike antigens in the cohorts to determine how they correlated with COVID-19 severity. The fold-change in HCoV response was calculated by dividing the antibody titres of each sample by the average titre in the SARS-CoV-2 seronegative blood donor cohort using the data show in Figure 1 (Figure 2a and Table S1, 18).

All cohorts previously infected with SARS-CoV-2 showed increased responses to the endemic beta-HCoV spike proteins relative to the unexposed background cohort (Figure 2a), suggesting that infection with SARS-CoV-2 induces increased cross-reactive responses, as reported elsewhere (10,12). No such trend was observed between the cohorts for antibody responses to alpha-HCoV spike proteins, which were comparable between all cohorts, irrespective of SARS-CoV-2 infection status.

We found that the increased reactivity to the beta-HCoV spike proteins was also broadly associated with COVID-19 severity. The response to the HCoV-OC43 spike antigen was significantly larger for those admitted to ICU than either the infected (p = 2.93×10^−6^) or asymptomatic groups (p = 3.73×10^−4^; Figure 3a). Similar increases were observed for the beta-HCoV HKU1 spike protein, although these were smaller in magnitude in comparison to those associated with the HCoV-OC43 spike protein (Figure 3a). As with antibody responses to the HCoV-OC43 spike protein, those who had to be treated in ICU had statistically significantly higher HCoV-HKU1 spike responses compared to asymptomatic (p = 9.2×10^−4^) and infected (p = 2.85×10^−7^) groups. However, significantly higher responses to the HCoV-HKU1 spike protein were not observed for those who were asymptomatic (p = 0.19) There was no significantly higher response to either the HCoV-OC43 (p = 0.07) or HCoV-HKU1 (p = 0.26) spike proteins in the SARS-CoV-2 antibody positive blood donor cohort.

**Figure 3:**
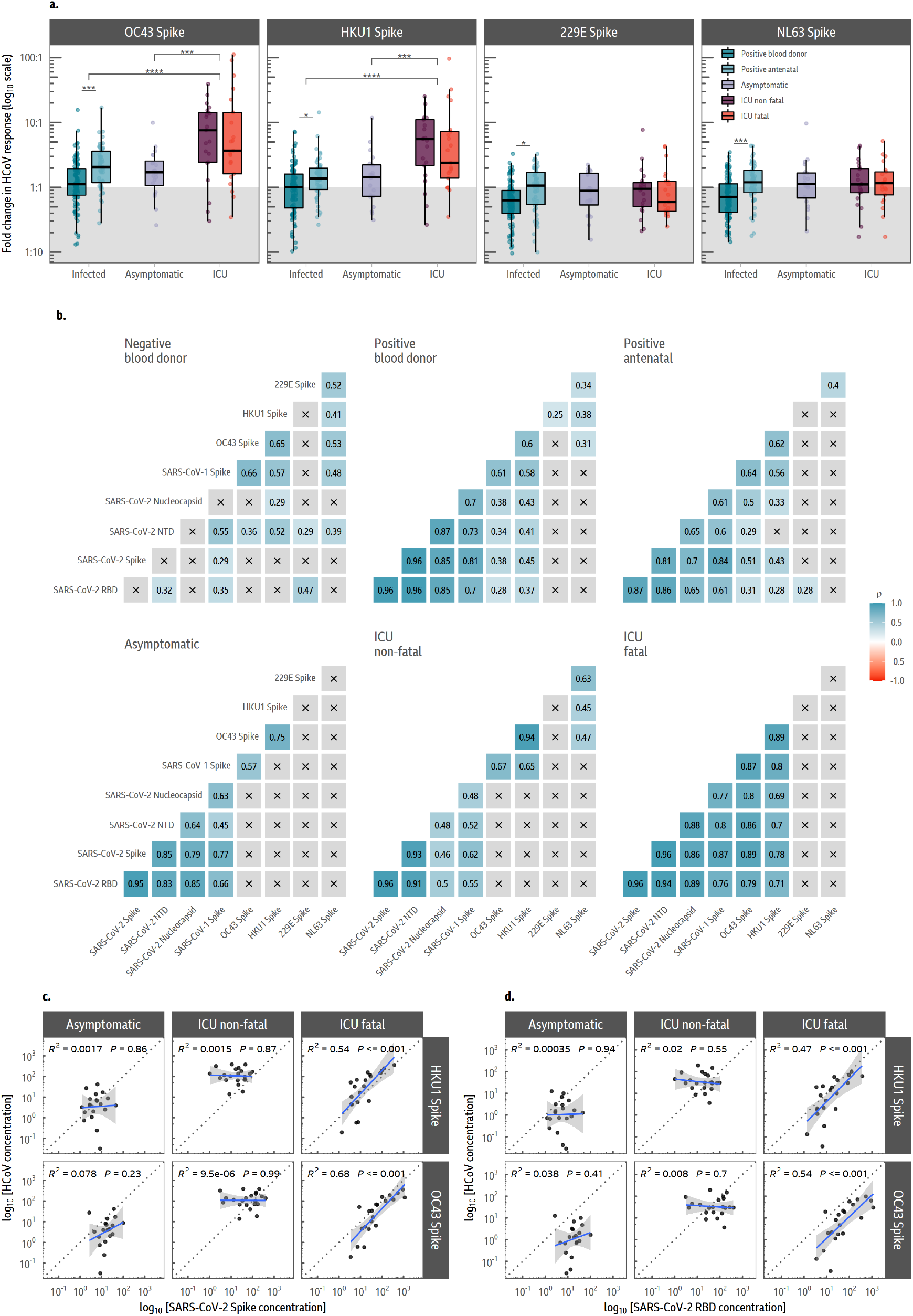
In fatal COVID-19, antibody responses to SARS-CoV-2 are highly correlated with antibody responses to the endemic beta-coronavirus spike proteins. **a. Mean fold-change of antibodies to endemic coronaviruses.** Antibodies to endemic beta-coronaviruses are boosted by SARS-CoV-2 infection and broadly correlate with COVID-19 severity. The mean fold-change to each spike protein was determined by dividing the average response to each antigen by responses in the background blood donor group (Table 1). Broad classifications of COVID-19 clinical presentations are given on the x-axis, with colour denoting sub-groups. A larger fold-change in OC43 and HCoV-HKU1 responses was observed in the ICU cohort relative to the blood donor, antenatal and asymptomatic groups, but not between infected and asymptomatic cohorts. The same increases were not observed for alpha-coronaviruses. There were no significant differences observed in the mean fold changes between the fatal and non-fatal ICU cohorts for any endemic HCoV. The data in this figure were generated using the ELISA-based MSD V-PLEX assay **b. Correlations between SARS-COV-2 and endemic coronavirus responses**. Spearman’s rank correlations (ρ) are shown for each pair of antigens, split by sample group. There is a positive correlation between all SARS-CoV-2 antigens in all but the background blood donor group. Significant correlations are found between SARS-CoV-2 epitopes and endemic beta-HCoVs (OC43 and HCoV-HKU1) in the positive blood donor, antenatal and fatal ICU groups; these correlations are absent in the asymptomatic and non-fatal ICU groups. The correlation between endemic beta-HCoVs and SARS-CoV-2 epitopes are considerably weaker in the larger positive blood donor and antenatal cohorts than in the fatal ICU samples. **c. and d. Responses to the SARS-CoV-2 spike (c) and receptor-binding domain (d) correlate with beta-coronavirus spike responses in fatal COVID-19**. Select correlations are shown in c. with a linear model fit between the concentration of two SARS-CoV-2 antigens and the endemic viruses OC43 and HCoV-HKU1. Data are shown only for sample groups with a clinical definition of COVID-19 severity (asymptomatic, non-fatal and fatal, across panel rows). The best fit line is shown in blue with 95% confidence intervals in grey; the dotted grey line denotes a 1:1 response to both antigens. There is a strong positive association between SARS-CoV-2 Spike/RBD and the endemic coronaviruses in the fatal ICU patients that is absent in the asymptomatic and non-fatal ICU patients.

There was no comparative increase in antibody responses to alpha-HCoV (HCoV-NL63 and HCoV-229E) spike proteins following SARS-CoV-2 infection in either the blood donor or asymptomatic cohorts. However, smaller increases in responses to alpha-HCoV spike protein were detected in the fatal/non-fatal ICU cohorts and the antenatal control group. For all endemic coronaviruses, the antenatal cohort had an elevated HCoV response in comparison to blood donors, which could be not explained by biases in age or sex, but we suggest these trends could be due to environmental differences (Figure 1; Figure 3a).

### Fatal COVID-19 outcomes are associated with a dominant immune response targeting shared beta-coronavirus epitopes

We then analysed how the SARS-CoV-2 antibody response correlated with the antibody response to beta-HCoV spike proteins. We calculated the Spearman’s rank correlation coefficients for all pairs of antigens from the V-PLEX assay, split by cohort (Figure 3b). Notably, in the fatal ICU cohort, responses to the SARS-CoV-2 spike were strongly correlated with the HCoV-OC43 and HCoV-HKU1 spike antigens (ρ = 0.89, p = 8×10^−8^ and ρ = 0.78, p = 4×10^−5^, respectively). Intriguingly, this correlation was present not only for the full-length spike antigen but also to the NTD (HCoV-OC43 p = 5×10^−7^; HCoV-HKU1 p = 4×10^−4^) and RBD (HCoV-OC43 p = 2×10^−5^; HCoV-HKU1 p = 3×10^−4^) of the spike, as well as the SARS-CoV-2 nucleocapsid (HCoV-OC43 p = 5.×10^−7^; HCoV-HKU1 p = 4×10^−4^; see Figure 3b). We could not identify statistically significant correlations in the similarly sized asymptomatic and non-fatal ICU cohorts. Similar correlations with beta-coronaviruses were also found for the SARS-CoV-2 spike S2 subunit (Figure S3).

A linear model fit on the log-scale was used to analyse the correlation of the magnitude of response to either the SARS-CoV-2 NTD or RBD of the spike, the full-length spike, and the SARS-CoV-2 nucleocapsid (Figures 3c & d, Figure S2 respectively) with the HCoV-HKU1 and HCoV-OC43 spike responses in the asymptomatic and ICU cohorts. Responses between SARS-CoV-2 antigens and the beta-HCoVs correlated strongly in the fatal ICU cohort with consistently high R^2^ values, indicating that for fatal COVID-19 outcomes the SARS-CoV-2 *de novo* antibody response is strongly linked with the responses to shared SARS-CoV-2/HCoV spike protein epitopes.

### Preferentially targeted epitopes map to the S2 subunit of the HCoV-OC43 spike protein

In fatal cases of COVID-19, responses to shared epitopes in the HCoV-OC43 spike protein increased in magnitude to a greater extent than those in the HCoV-HKU1 spike protein (Figure 3). To identify the location of the beta-HCoV response causing the correlation, we subdivided the HCoV-OC43 spike protein into the NTD (aa 1-419) as well as the S1 (aa 1-794) and S2 (aa 766-1304) subunits of the spike protein (Figure 4a).

**Figure 4:**
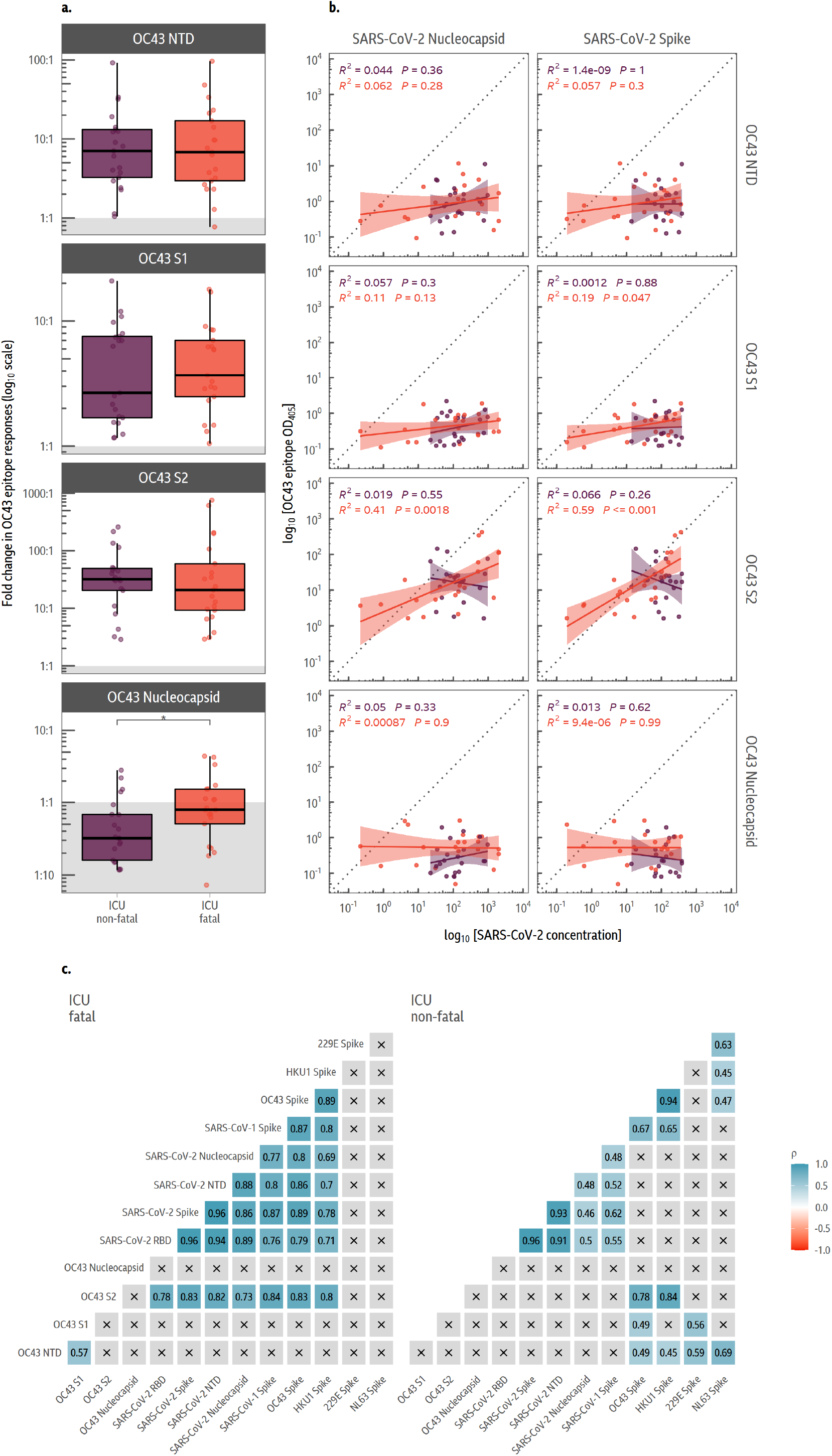
Antibody responses are directed against the S2 subunit of the HCoV-OC43 spike protein. **a. Fold-change in responses to various domains/subunits in the HCoV-OC43 spike protein and nucleocapsid.** Indirect ELISAs were run against the N-terminal domain (NTD), S1 subunit and S2 subunit of the HCoV-OC43 spike protein, in addition to the HCoV-OC43 nucleocapsid. Fold-change via ELISA was determined relative to the average value in the SARS-CoV-2 negative blood donor cohort. Antibody levels are increased against all antigens apart from the nucleocapsid, with the largest increase in antibody response to the S2 subunit of the spike protein. **b. Correlation in responses between SARS-CoV-2 antigens and HCoV-OC43 spike protein domains and nucleocapsid**. The log-scale OD_405_ values from the HCoV-OC43 spike and nucleocapsid ELISA (along the rows) is compared to the MSD V-PLEX SARS-CoV-2 results (columns). A linear model is fit on the log-scale is annotated with the associated 95% confidence intervals, R^2^ and p-value. Values and model fits for the non-fatal ICU samples is given in purple, while red is used for the fatal group. The HCoV-OC43 S2 subunit ELISA result is correlated with the concentration of SARS-CoV-2 antibodies, but only in the fatal group. **c. Correlations between ELISAs and MSD V-PLEX SARS-CoV-2 assay responses**. Responses to the S2 subunit of HCoV-OC43 are strongly correlated with the MSD concentration of SARS-CoV-2 antibodies in those who died from coronavirus, but not those who survived. Notably, there is a positive correlation between the S2 subunit response and the HCoV-OC43 Spike and HCoV-HKU1 concentrations in both cohorts, suggesting that this is a shared antibody target among the endemic beta-coronaviruses.

In response to both fatal and non-fatal SARS-CoV-2 infection, there was an increase in response to all the HCoV-OC43 spike domains analysed, although the response to the S2 subunit was considerably greater, indicating that the majority of shared SARS-CoV-2 and beta-HCoV epitopes reside in the S2 subunit.

There were no significant differences in the fold-change of responses to these specific regions between ICU patients with non-fatal and fatal COVID-19 outcomes (Figure 3b). There were median fold-increases of 6.93 (p = 1×10^−3^), 2.48 (p = 4×10^−3^), 31.4 (p = 2.×10^−18^) increases to the NTD, S1 and S2 HCoV-OC43 spike domains, respectively, across the fatal and non-fatal ICU cohorts in comparison to the blood donor control group.

We fitted a linear regression between the log-concentration of response between the HCoV-OC43 responses (NTD, S1, S2 and nucelocapsid) and either the SARS-CoV-2 full-length spike protein or nucleocapsid (Figure 4b). In those individuals with fatal COVID-19 outcomes, there was a strong correlation between antibody responses to the HCoV-OC43 S2 subunit and the SARS-CoV-2 spike, which extended to the SARS-CoV-2 RBD and NTD of the spike protein, as well as the SARS-CoV-2 nucleocapsid (Figure 4c). The SARS- CoV-2 spike S2 subunit has previously been shown to be the target of antibodies induced by prior endemic coronavirus infection (9,10; Figure 4c). In contrast to the S2 spike subunit, antibody responses to both the HCoV-OC43 S1 subunit and NTD correlated poorly with SARS-CoV-2 antibody response in fatal COVID-19 outcomes (Figure 4c).

We then compared the response to the HCoV-OC43 nucleocapsid in individuals with fatal and non-fatal COVID-19 outcomes. Whilst the response to the SARS-CoV-2 spike was lower in those with fatal disease, the SARS-CoV-2 nucleocapsid response was not (Figure 1).

In comparison to the SARS-CoV-2 spike response where responses to the HCoV-OC43 increased upon SARS-CoV-2 infection, no increase in antibody responses to the HCoV-OC43 nucleocapsid were detected upon SARS-CoV-2 infection (Figure 4a). The non-fatal cohort had a median fold change of 0.79 (p = 4×10^−4^), whilst there was no change for the fatal cohort when compared to the control group (p = 0.28). The absence of any increased cross-reactivity to the HCoV-OC43 nucleocapsid in individuals infected with SARS-CoV-2 indicates that unlike in the case of the SARS-CoV-2 spike, the SARS-CoV-2 immune response was not targeting epitopes shared between the HCoV-OC43 and SARS-CoV-2 nucleocapsids (Figure 4). Consequently, the nucleocapsid *de novo* response appears not to be linked with the responses to shared SARS-CoV-2/beta-HCoV epitopes, unlike that of the SARS-CoV-2 spike protein.

## Discussion

Our study shows that a dominant immune response targeting shared beta-coronavirus epitopes in the S2 subunit of the spike protein is associated with fatal COVID-19 disease. To date, this is the first study to associate previous exposure to HCoVs directly with COVID-19 clinical outcomes.

Our results are in line with previous work (22) showing higher neutralising antibody titres and increased antibody levels to a range of beta-coronavirus antigens in patients with severe COVID-19. However, in contrast to those studies, we found that titres were significantly lower among patients with a fatal COVID-19 outcome. There was also a strong correlation in the cohort with a fatal COVID-19 outcome between antibody responses to SARS-CoV-2 spike and nucleocapsid, and antibody responses to both HCoV-HKU1 and HCoV-OC43 spike proteins. This correlation was shown to be due to preferential targeting of the HCoV-OC43 S2 subunit, but not the HCoV-OC43 NTD or S1 subunit (Figure 3; 9, 10).

These observations suggest that individuals with fatal COVID-19 outcomes are failing to mount the same level of *de novo* immune responses to novel epitopes specific to the SARS-CoV-2 spike as those who had non-fatal outcomes. This could reflect an immune response that is influenced by past exposure to endemic beta-coronaviruses, likely due to the recall of memory B cells dominating the SARS-CoV-2 response (23) abrogating or diverting an effective new immune response to a new SARS-CoV-2 infection.

This hypothesis is supported by (i) previous publications demonstrating that the majority of detectable cross-reactive antibody response induced by prior HCoV infection are directed at the S2 subunit of the SARS-CoV-2 spike (9,10), (ii) the strong correlation between the antibody response to the S2 subunit of the HCoV-OC43 spike induced by SARS-CoV-2 infection and the response to the SARS-CoV-2 full-length spike, NTD, RBD and the nucleocapsid in fatal COVID-19, and (iii) the inhibition of responses to the RBD, NTD and full-length spike, but not the S2 subunit or SARS-CoV-2 nucleocapsid during SARS-CoV-2 infection.

This mechanism may be reminiscent of original antigenic sin (OAS) as it involves prior immune responses compromising the generation of *de novo* responses to a closely related pathogen (24). Similar phenomena have been observed for dengue and influenza viruses (25,26). However, it should be considered that the examples documented to date relate to different variants or serotypes of the same pathogen.

The inhibition in the development of *de novo* responses to new SARS-CoV-2 epitopes does not appear to extend to the SARS-CoV-2 nucleocapsid protein. It seems that this is due to the absence of targeted shared epitopes between the SARS-CoV-2 and endemic coronavirus nucleocapsids allowing sufficient recall of memory B cells to compromise the development of responses to novel nucleoprotein protein epitopes (Figure 3). The absence of shared SARS-CoV-2/HCoV-OC43 nucleocapsid epitopes was demonstrated by the absence of any increase in response to the HCoV-OC43 nucleocapsid upon SARS-CoV-2 infection.

The timeframe of sampling has been suggested to be critical in the ability to identify differences in the responses amongst severely ill COVID-19 patients (27). The single timepoint sampled in this study limits the window in which the appearance of *de novo* responses can be examined in fatal COVID-19 cases. Consequently, an earlier timepoint might indicate that neutralising antibodies are generally lower, as opposed to only one out of three neutralisation or ACE2 binding assays showing this feature (Figure 2). This would match with the consistently lower IgG RBD, NTD and spike responses. Our data proposes a probable mechanism for this feature – a dominant immune response directed against shared beta-HCoV epitopes in the S2 subunit of the SARS-CoV-2 spike.

This study is further limited by the absence of longitudinal samples, which might help determine why some individuals mount a targeted S2 response whilst others do not. Our HCoV-OC43 nucleocapsid protein assay shows this is not due to recent HCoV-OC43 infection, which would have shown increased responses to HCoV-OC43 nucleocapsid relative to the negative blood donor controls (Figure 3c). The correlation between fatal COVID-19 and the observed targeted antibody response is likely due to an individual’s past history of infection with HCoV-OC43/-HKU1. However, whether the key contributing factors are age, sex, time, since the last beta-coronavirus infection, infection with specific strains or just serendipity is beyond the scope of this study.

## Data Availability

Data is available on request and will be submitted into a suitable repository upon acceptance of the paper.

## Materials and Methods

### Study Approval

Ethical approval was obtained for the Scottish National Blood Transfusion Service (SNBTS) anonymous archive - IRAS project number 18005. SNBTS blood donors gave fully informed consent to virological testing, donation was made under the SNBTS Blood Establishment Authorisation and the study was approved by the SNBTS Research and Sample Governance Committee IRAS project number 18005.

The ISARIC WHO CCP-UK protocol was developed by international consensus in 2012-14 and activated in response to the SARS-CoV-2 pandemic on 17th January 2020. This is an actively recruiting prospective cohort study recruiting across the United Kingdom (28). Study materials including protocol, revision history, case report forms, study information and consent forms, are available online [https://isaric4c.net/protocols/]. Ethical approval was given by the South Central - Oxford C Research Ethics Committee in England (Ref: 13/SC/0149) and by the Scotland A Research Ethics Committee (Ref: 20/SS/0028).

The antenatal samples were collected during routine antenatal care appointments in the Oxfordshire area. Samples were taken during the first trimester of pregnancy (generally between 8–12 weeks’ gestation) between 14 April and 15 June 2020. Ethical approval was obtained from the South-Central Research Ethics Committee (Ref: 08/H0606/139).

Patients and healthcare workers comprising the asymptomatic cohort were recruited from the John Radcliffe Hospital in Oxford, United Kingdom, between March and May 2020. Patients identified during the SARS-COV-2 pandemic were screened and recruited into the Sepsis Immunomics (IRAS260007) and International Severe Acute Respiratory and Emerging Infection Consortium World Health Organization Clinical Characterisation Protocol UK (IRAS126600). Patients were sampled at least 28 days from their positive RT-PCR test. The ICU patients were enrolled as part of an ongoing prospective observational study AspiFlu (<u>ISRCTN51287266</u>) at St George’s Hospital, London, UK. Researchers working with the samples in the laboratory were blinded to the clinical outcomes of the ICU patients during testing. None of the study subjects received convalescent plasma therapy.

### Enzyme-linked immunosorbent assay

SARS-CoV-2 spike, RBD as well as HCoV-229E, HCoV-NL63, HCoV-HKU1 and HCoV-OC43 spike IgG antibody responses were measured using ELISAs. Further work to characterise the location of the conserved epitopes between HCoV-OC43 and SARS-CoV-2 used the HCoV-OC43, NTD, S1 and S2 subunits. Spike and RBD proteins were produced as per Armanat et al 2020 (29). HCoV-229E, HCoV-NL63, HCoV-HKU1 and HCoV-OC43 spike antigens were bought from Sino Biological, China. The S1 and S2 proteins were bought from Sino Biological, China an the OC43 NTD from The Native Antigen, UK.

Nunc-Immuno 96-well plates (Thermo Fischer Scientific, USA) were coated with 1.0 μg ml^−1^ of antigen in PBS buffer and left overnight at 4 °C. Plates were washed with 3x with 0.1% PBS–Tween (PBS/T), then blocked with casein in PBS for 1 hour at room temperature (RT). Serum or plasma was diluted in casein–PBS solution at dilutions ranging from 1:50 to 1:20,000 before being added to Nunc-Immuno 96-well plates in triplicate. Plates were incubated for 2 hours before being washed with 6x with PBS/T. Secondary antibody rabbit anti-human whole IgG conjugated to alkaline phosphatase (Sigma, USA) was added at a dilution of 1:1000 in casein–PBS solution and incubated for 1 hour at RT. After a final wash, plates were developed by adding 4-nitrophenyl phosphate substrate in diethanolamine buffer (Pierce, Loughborough, UK), and optical density (OD) was read at 405 nm using an ELx800 microplate reader (Cole Parmer, London, UK). A reference standard comprising of pooled cross-reactive serum and naïve serum on each plate served as positive and negative controls, respectively.

The positive reference standard was used on each plate to produce a standard curve. A monoclonal antibody standard was used for the RBD/spike ELISAs. Pooled HCoV highly reactive sera were used as a standard for the HCoV spike ELISAs.

### Protein sequences used in ELISAs

| Protein | Expressed in | Provider | Reference (if appropriate) |
| --- | --- | --- | --- |
| HCoV-HKU1 spike | Insect cells | Sino Biological |  |
| HCoV-OC43 spike | Insect cells | Sino Biological |  |
| HCoV-NL63 spike | Insect cells | Sino Biological |  |
| HCoV-229E spike | Insect cells | Sino Biological |  |
| SARS-CoV-2 spike | HEK 293T | Produced in-house | Amanat et al 2020 (29) |
| SARS-CoV-2 RBD | HEK 293T | Produced in-house | Amanat et al 2020 (29) |
| SARS-CoV-2 S2 subunit | HEK 293T | Sino Biological |  |
| HCoV-OC43 S1 subunit | HEK 293T | Sino Biological |  |
| HCoV-OC43 S2 subunit | Insect cells | Sino Biological |  |
| HCoV-OC43 NTD | *Escherichia coli* | The Native Antigen Company |  |

### MSD V-PLEX assay

IgG antibody responses to SARS-CoV-2 spike, RBD, NTD and nucleocapsid and the spike proteins of SARS-CoV-1, HCoV-229E, HCoV-NL63, HCoV-HKU1 and HCoV-OC43 were assessed using the Meso Scale Diagnostics (MSD) Multi-Spot Assay System (MSD, USA). Pre-coated plates (‘Coronavirus panel 2’) were incubated at room temperature (RT) with Blocker A solution for at least 30 minutes whilst being shaken at 500-700 rpm. Serum or plasma was diluted in Diluent 100 at dilutions of 1:500 to 1:50,000 and samples were added to the plates in duplicate. Plates were incubated for 2 hours at RT, whilst being shaken at 500-700 rpm throughout. A 1x working concentration of the SULFO-TAG anti-human IgG Detection Antibody was prepared in Diluent 100. After incubation with the samples, the plates were washed x3 with 1x MSD Wash buffer. Prepared detection antibody solution was added to the plates, which were incubated at RT for 1 hour, whilst being shaken. Plates were then washed x3 with 1X MSD Wash buffer. To read the assay results, MSD GOLD Read Buffer B (provided ready to use) was added to the plate. No incubation is required, and the plates were read on a MESO QuickPlex SQ 120 (MSD, USA) immediately after adding the buffer.

A 7-point calibration curve of the standards was prepared using Diluent 100. Diluent 100 was used as a negative control. An additional three positive controls provided with the kit were also run on every plate. All standards and controls were run in duplicate. Data from the assay was analysed using MSD Discovery Workbench software, which averaged all the duplicates, generated and fitted all the data to standard curves.

### MSD ACE2 competition assay

The ability of antibodies present in serum/plasma to inhibit the binding of angiotensin-converting enzyme 2 (ACE2) to the SARS-CoV full-length spike proteins and RBD domains was assessed using the COVID-19 ACE2 competition assay (MSD, USA). The assay can be used to estimate the neutralising activity of the antibodies present in the samples.

Pre-coated plates were incubated at RT with Blocker A solution for at least 30 minutes whilst being shaken at 500-700 rpm. Serum or plasma was diluted in Diluent 100 at dilutions of 1:10 to 1:100 and samples were added to the plates in duplicate. Plates were incubated for 1 hour at RT, whilst being shaken at 500-700 rpm throughout. A 1x working concentration of the SULFO-TAG ACE2 detect was prepared in Diluent 100. After incubation with the samples, the plates were washed x3 with f 1x MSD Wash buffer. Prepared SULFO-TAG ACE2 solution was added to the plates, which were incubated at RT for a further 1 hour, whilst being shaken. Plates were then washed x3 with 1X MSD Wash buffer. To read the assay results, MSD GOLD Read Buffer B (provided ready to use) was added to the plate. Plates were read immediately after adding the buffer on a MESO QuickPlex SQ 120 (MSD, USA)

A 7-point calibration curve of the standards was prepared using Diluent 100. Diluent 100 was used as a negative control. All standards were run in duplicate. Data from the assay was analysed using MSD Discovery Workbench software, which averaged all the duplicates, generated and fitted all the data to standard curves.

### Pseudotyped virus microneutralisation assay

A lentivirus-based pseudotyped virus system was used to display the SARS-CoV-2 spike protein on its surface using a synthetic codon optimised SARS-CoV-2 expression construct (NCBI reference sequence: YP_009724390.1).

Target cells were transfected 24 hours prior to assay setup with 2.75 ug of ACE2 expression plasmid and 250 ng of TPRSSM2 expression plasmid (20).

### Data analysis

Over dispersed variables were transformed onto the logarithmic scale (base 10) for between group comparisons for V-PLEX platform concentrations, ELISA optical density and neutralising titres. Unless otherwise specified, a t-test assuming unequal variances was used to test for differences in the mean responses; values were analysed on the logarithmic (base 10) scale unless otherwise stated. A Holm correction was applied to p-values for multiple comparisons. In cases where a fold change or ratio is calculated, the log-scale group means can be compared to zero using a t-test to determine if the group differs from equal concentrations of antigens. For non-normal variables that do not meet the assumptions of normality – even if transformed – a Dunn’s test was used instead. Reported correlations are Spearman’s rank, as the measure is non-parametric and robust to transformation.

## Author Contributions

Conceptualisation, PK, SG, CPT; Methodology, ALM, RSP, ME, PP, KK, NT, CW, SR, TL, KK; Software Programming, RSP, JB, UO, JL; Formal Analysis, RSP, CPT, KK; Investigation, ALM, ME, AF, JW, HK, JB, JR, LL, LS; Resources, SB-R, GC, WD, CD, DWE, AH, SL, CL-C, SFL, DM, AJM, NP, DS, PS, NT, CW, BW, DW, KK, MGS, JKB, PJMO ISARIC4C Investigators, OPTIC consortium, SBTS consortium, PS, MGS, TL, KB, MT, MC, GS, SHK, LJ EB, SD, PCM, TB; Data Curation Management, ALM, RSP, ME, JY, JW, SB-R, GC, WD, CD, DWE, AH, SL, CL-C, SFL, DM, AJM, NP, DS, PS, NT, CW, BW, DW, ISARIC4C Investigators, OPTIC consortium, SBTS consortium; Writing – Original Draft Preparation, ALM, RSP, CPT; Writing – Review & Editing Preparation, all authors reviewed and approved the final manuscript; Visualization Preparation, RSP, UO, JL, CPT; Supervision, TL, SD, PK, SG, CT; Project Administration, ALM, CPT; Funding Acquisition, PK, SG, CPT, MGS, JKB, PJMO.

## Acknowledgments

We would like to express our gratitude and acknowledge the contribution of the staff at Oxford University Hospitals NHS Foundation Trust, Scottish National Blood Transfusion Service and St George’s University Hospitals NHS Foundation Trust (London) who were involved in the provision and preparation of samples analysed in this project. We acknowledge the wider support of the ISARIC4C, OPTIC and SBTS research consortia.

This work was supported by the Georg and Emily von Opel Foundation, the National Institute for Health Research (NIHR) [award CO-CIN-01], the Medical Research Council [grant MC_PC_19059], ME was supported by The Leona M. and Harry B. Helmsley Charitable Trust on May 31, 2021 for the project titled, “ICARUS –IBD: International study of COVID-19 Antibody Response Under Sustained immune suppression in Inflammatory Bowel Disease. RP also supported by funds provided under Professor RW Snow’s Wellcome Trust Principal Fellowship (# 212176). Meso Scale Diagnostics (USA) provided loan of equipment, reagents and technical support. JSB was supported by funding from the Biotechnology and Biological Sciences Research Council (BBSRC) [grant number BB/M011224/1]. CPT was funded by an ERC research grant ‘UNIFLUVAC’ and two MRC CiC grants (Ref: BR00140). ALM is funded by a NIHR Research Capability Funding grant. HJK is supported by The Future of Humanity Institute at the University of Oxford DPhil Scholarship program. DE is funded by the Robertson Foundation. The funders played no role in the design, execution or reporting of the study.

## Competing interests

DE declares lecture fees from Gilead. CPT and SG hold funding from Blue Water Vaccines.

## Supplementary Material

**Figure S1:**
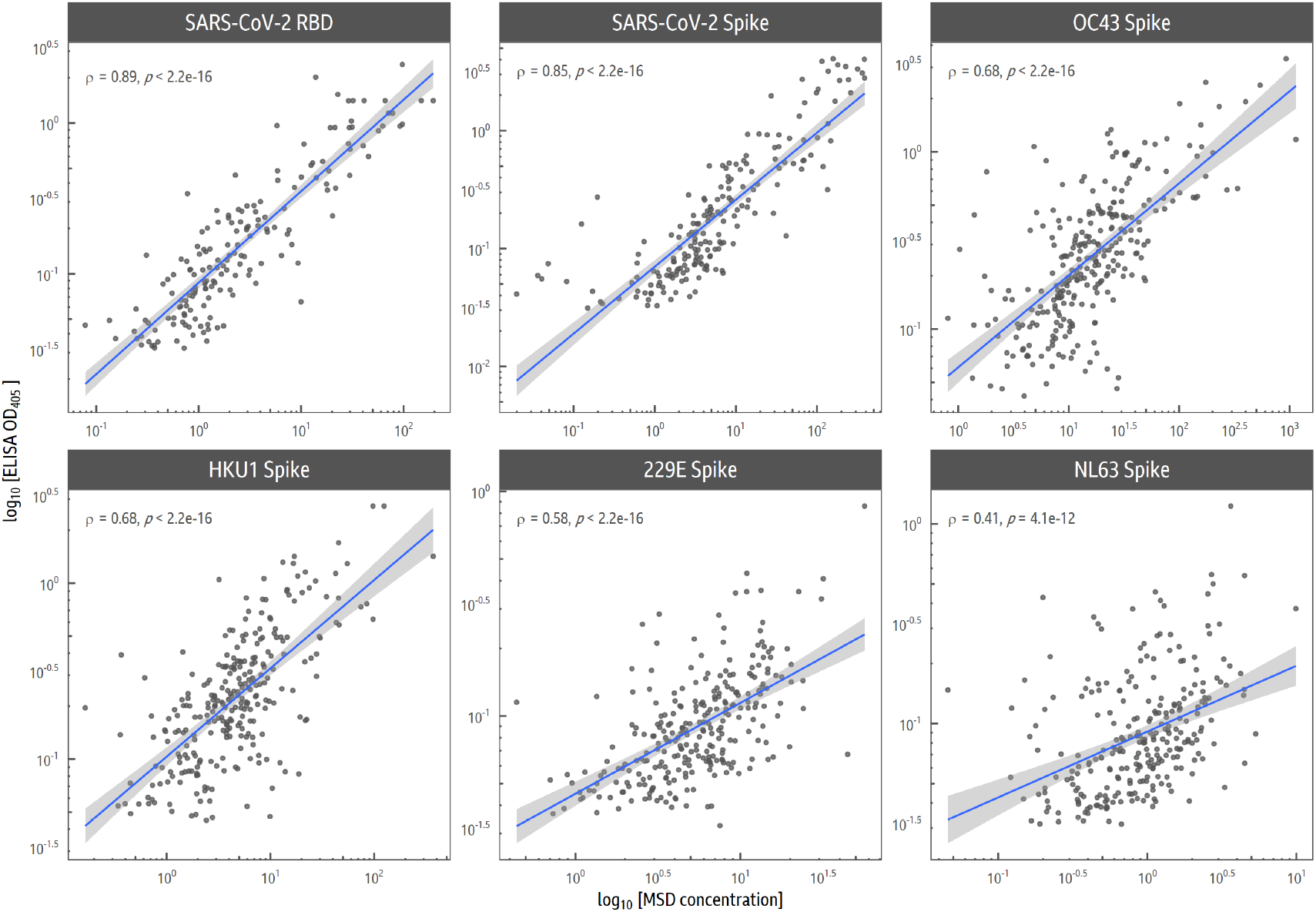
Correlation of MSD VPLEX and ELISA data. The correlation of results across the two assays for the SARS-CoV-2 full-length spike and RBD domains, along with endemic coronavirus spike antigens is shown, with Spearman Rank correlations.

**Figure S2:**
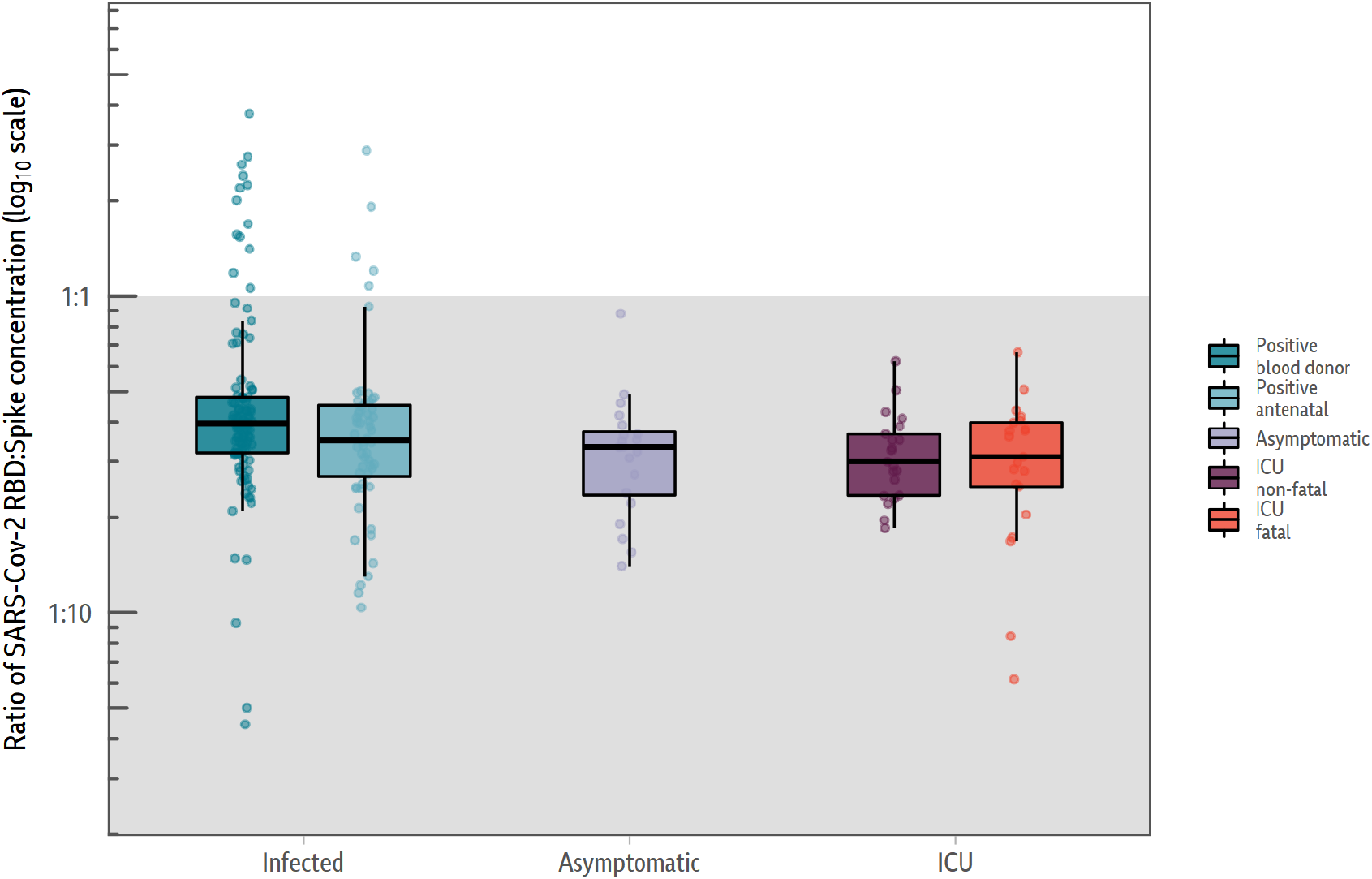
Ratio of SARS-CoV-2 RBD response to spike response as measured by MSD VPLEX assay.

**Figure S3:**
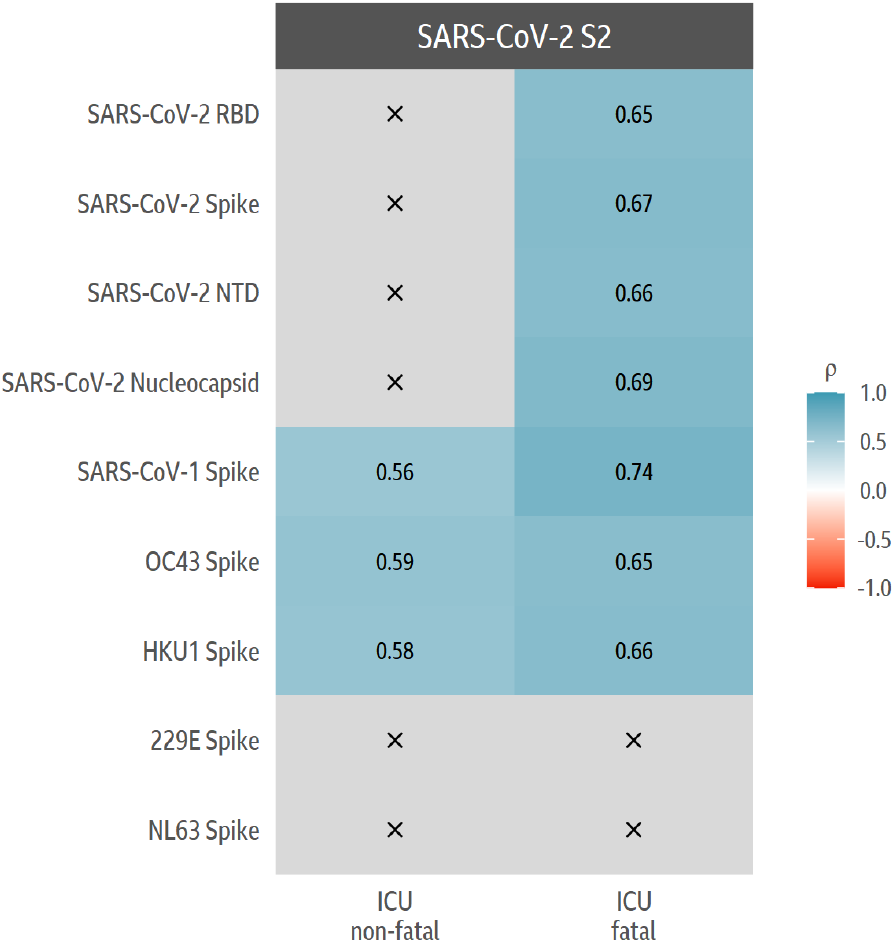
Correlations of the S2 ELISA antibody responses in fatal and non-fatal individuals with SARS-CoV-2 and HCoV antigens.

**Figure S4:**
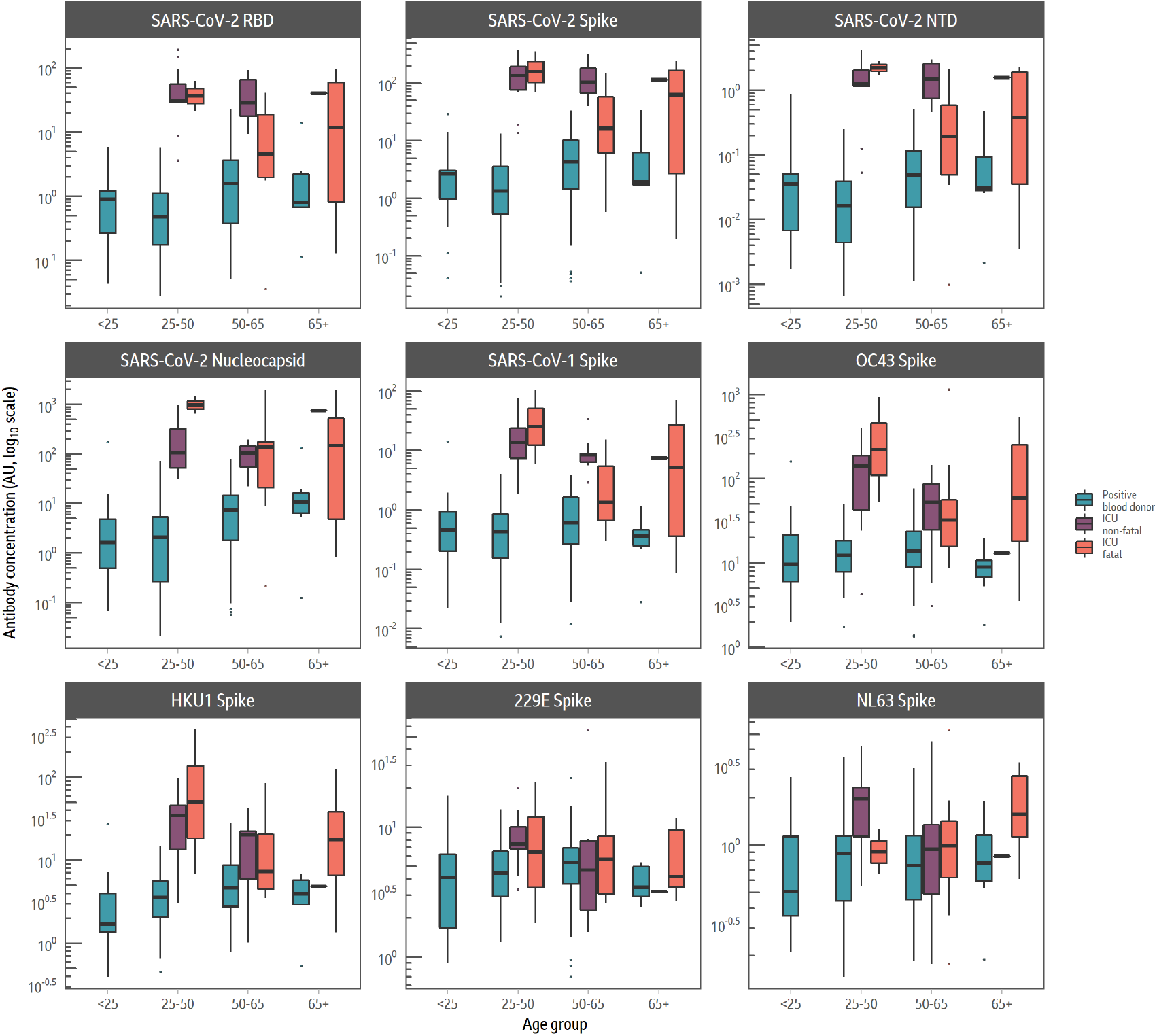
Responses categorised by age in the different groups.

**Figure S5:**
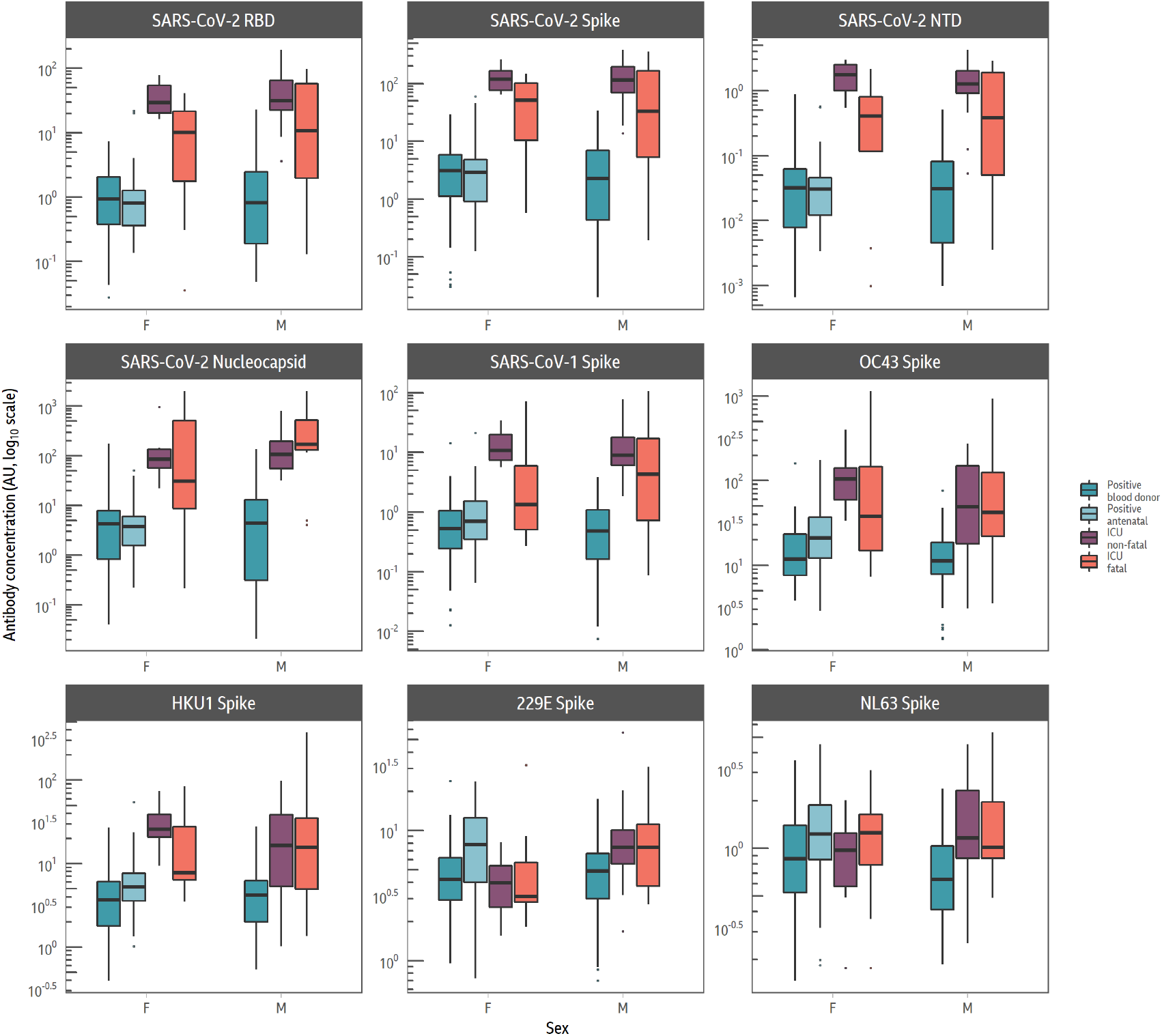
Responses categorised by sex in the different groups.

**S6:**
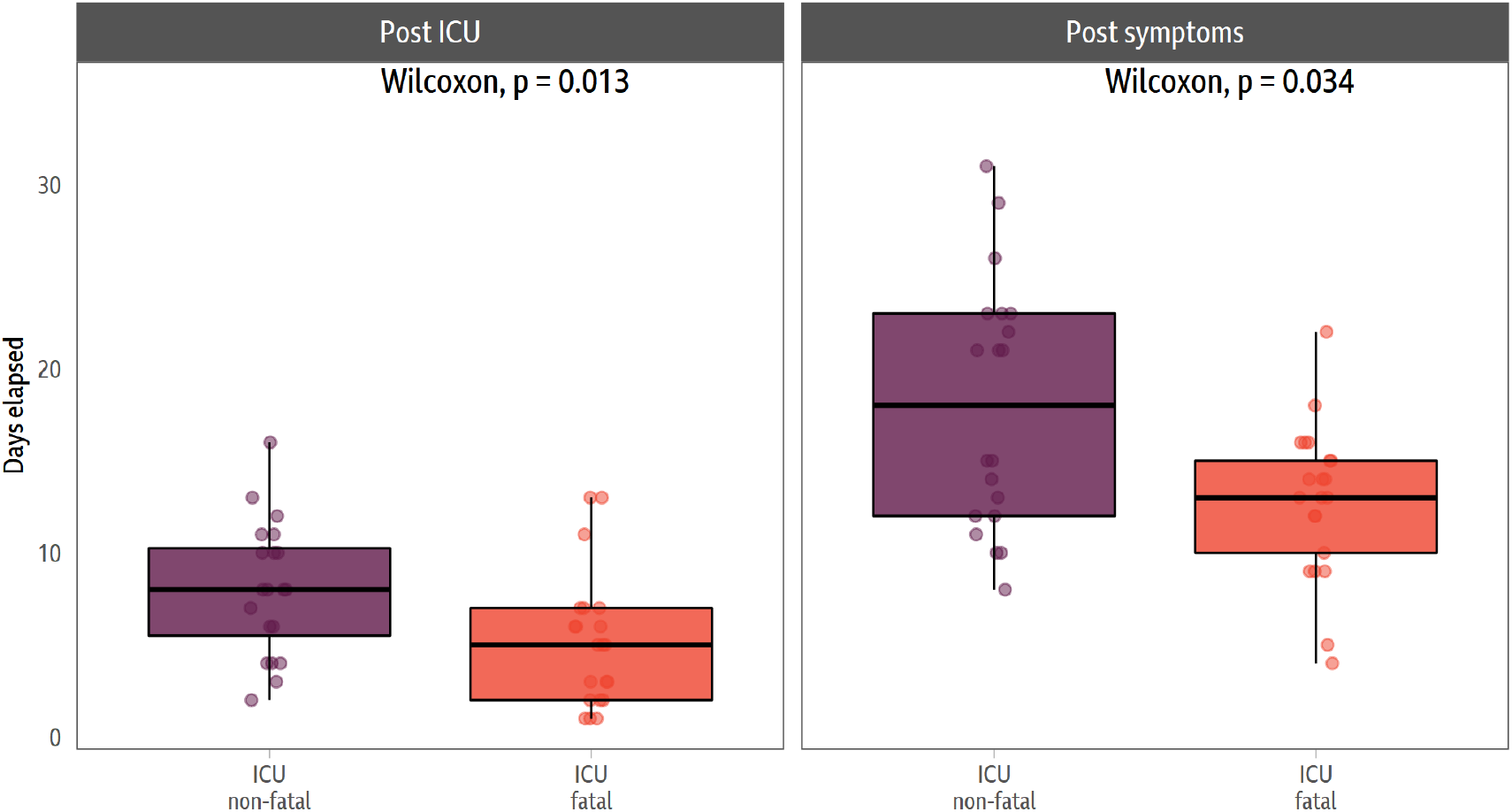
Days from post-symptomatic and entry to ICU in the fatal and non-fatal groups. In both instances there is statistically significant differences.

## Notes

### Author Declarations

Blood donor samples: The study was approved by the SNBTS Research and Sample Governance Committee IRAS project number 18005. Antenatal samples: Ethical approval was obtained from the South-Central Research Ethics Committee (Ref: 08/H0606/139). 14 of the 22 asymptomatic samples: Ethical approval was given by the South Central - Oxford C Research Ethics Committee in England (Ref: 13/SC/0149) and by the Scotland A Research Ethics Committee (Ref: 20/SS/0028). 8 of the 22 asymptomatic samples: Samples from healthcare workers who had asymptomatic SARS-CoV-2 infection were collected as part of the Oxford Translational Gastrointestinal Unit GI Biobank COVID Study 16/YH/0247 [research ethics committee (REC) at Yorkshire & The Humber : Sheffield]. ICU samples (both fatal and non-fatal cohorts) The ICU patients were enrolled as part of an on going prospective observational study AspiFlu (ISRCTN51287266) which has national HRA (CPMS 43440/IRAS 271269) and REC (19/WA/0310) approval.

## References

1. Gaunt ER, Hardie A, Claas ECJ, Simmonds P, Templeton KE. Epidemiology and clinical presentations of the four human coronaviruses 229E, HKU1, NL63, and OC43 detected over 3 years using a novel multiplex real--time PCR method. J Clin Microbiol. 2010;48(8):2940–7.

2. Petrosillo N, Viceconte G, Ergonul O, Ippolito G, Petersen E. COVID--19, SARS and MERS: are they closely related? Clinical Microbiology and Infection. 2020. p. 729–34.

3. Li Q, Guan X, Wu P, Wang X, Zhou L, Tong Y, et al. Early transmission dynamics in Wuhan, China, of novel coronavirus--infected pneumonia. New England Journal of Medicine. 2020. p. 1199–207.

4. Bacher P, Rosati E, Esser D, Martini GR, Saggau C, Schiminsky E, et al. Low avidity CD4+ T cell responses to SARS--CoV--2 in unexposed individuals and humans with severe COVID--19. Immunity [Internet]. Elsevier Inc.; 2020; Available from: https://doi.org/10.1016/j.immuni.2020.11.016

5. Mateus J, Grifoni A, Tarke A, Sidney J, Ramirez SI, Dan JM, et al. Selective and cross--reactive SARS--CoV--2 T cell epitopes in unexposed humans. Science (80--). 2020;370(6512).

6. Braun J, Loyal L, Frentsch M, Wendisch D, Georg P, Kurth F, et al. SARS--CoV--2--reactive T cells in healthy donors and patients with COVID--19. Nature. 2020;587(7833):270–4.

7. Le Bert N, Tan AT, Kunasegaran K, Tham CYL, Hafezi M, Chia A, et al. SARS--CoV--2--specific T cell immunity in cases of COVID--19 and SARS, and uninfected controls. Nature. 2020;584(7821):457–62.

8. Lipsitch M, Grad YH, Sette A, Crotty S. Cross--reactive memory T cells and herd immunity to SARS--CoV--2. Nature Reviews Immunology. 2020. p. 709–13.

9. Ng KW, Faulkner N, Cornish GH, Rosa A, Harvey R, Hussain S, et al. Preexisting and de novo humoral immunity to SARS--CoV--2 in humans. Science (80--). 2020;eabe1107.

10. Anderson EM, Goodwin EC, Verma A, Arevalo CP, Bolton MJ, Weirick ME, et al. Seasonal human coronavirus antibodies are boosted upon SARS--CoV--2 infection but not associated with protection. Cell. 2021;

11. Addetia A, Crawford KHD, Dingens A, Zhu H, Roychoudhury P, Huang ML, et al. Neutralizing antibodies correlate with protection from SARS--CoV--2 in humans during a fishery vessel outbreak with a high attack rate. J Clin Microbiol. 2020;58(11).

12. Westerhuis BM, de Bruin E, Chandler FD, Ramakers CRB, Okba NMA, Li W, et al. Homologous and heterologous antibodies to coronavirus 229E, NL63, OC43, HKU1, SARS, MERS and SARS--CoV--2 antigens in an age stratified cross--sectional serosurvey in a large tertiary hospital in The Netherlands. medRxiv [Internet]. 2020 Jan 1;2020.08.21.20177857. Available from: http://medrxiv.org/content/early/2020/08/24/2020.08.21.20177857.abstract

13. Ng K, Faulkner N, Cornish G, Rosa A, Earl C, Wrobel A, et al. Pre--existing and de novo humoral immunity to SARS--CoV--2 in humans. bioRxiv. 2020;2020.05.14.095414.

14. Westerhuis BM, Aguilar--Bretones M, Raadsen MP, de Bruin E, Okba NMA, Haagmans BL, et al. Severe COVID--19 patients display a back boost of seasonal coronavirus--specific antibodies. medRxiv [Internet]. 2020 Jan 1;2020.10.10.20210070. Available from: http://medrxiv.org/content/early/2020/10/12/2020.10.10.20210070.abstract

15. Aydillo T, Rombauts A, Stadlbauer D, Aslam S, Abelenda--Alonso G, Escalera A, et al. Antibody Immunological Imprinting on COVID--19 Patients. medRxiv [Internet]. 2020;2020.10.14.20212662. Available from: https://doi.org/10.1101/2020.10.14.20212662

16. Weiskopf D, Schmitz KS, Raadsen MP, Grifoni A, Okba NMA, Endeman H, et al. Phenotype and kinetics of SARS--CoV--2--specific T cells in COVID--19 patients with acute respiratory distress syndrome. Sci Immunol. 2020;5(48).

17. Ogbe A, Kronsteiner B, Skelly DT, Pace M, et al. T cell assays differentiate clinical and subclinical SARS--CoV--2 infections from cross--reactive antiviral responses. Nat Commun. 2021;12(1).

18. Carvalho T. Cross--reactive T cells. Nature Medicine. 2020. p. 1807.

19. Lumley SF, Eyre DW, McNaughton AL, Howarth A, Hoosdally S, Hatch SB, et al. SARS--CoV--2 antibody prevalence, titres and neutralising activity in an antenatal cohort, United Kingdom, 14 April to 15 June 2020. Euro Surveill. 2020;25(42).

20. Thompson CP, Grayson NE, Paton RS, Bolton JS, Lourenço J, Penman BS, et al. Detection of neutralising antibodies to SARS--CoV--2 to determine population exposure in Scottish blood donors between March and May 2020. Eurosurveillance. 2020;25(42).

21. Kooblall KG, Boon H, Cranston T, Stevenson M, Pagnamenta AT, Rogers A, et al. Multiple Endocrine Neoplasia Type 1 (MEN1) 5′UTR Deletion, in MEN1 Family, Decreases Menin Expression. J Bone Miner Res. 2021;36(1):100–9.

22. Seow J, Graham C, Merrick B, Acors S, Pickering S, Steel KJA, et al. Longitudinal observation and decline of neutralizing antibody responses in the three months following SARS--CoV--2 infection in humans. Nat Microbiol. 2020;5(12):1598–607.

23. Wong R, Bhattacharya D. Basics of memory B--cell responses: lessons from and for the real world. Immunology. 2019. p. 120–9.

24. Francis TJ. On the Doctrine of Original Antigenic Sin. Proc Am Philos Soc [Internet]. 1960;104(6):572–8. Available from: http://www.jstor.org/stable/985534

25. Mongkolsapaya J, Dejnirattisai W, Xu XN, Vasanawathana S, Tangthawornchaikul N, Chairunsri A, et al. Original antigenic sin and apoptosis in the pathogenesis of dengue hemorrhagic fever. Nat Med. 2003;9(7):921–7.

26. Linderman SL, Chambers BS, Zost SJ, Parkhouse K, Li Y, Herrmann C, et al. Potential antigenic explanation for atypical H1N1 infections among middle--aged adults during the 2013--2014 influenza season. Proc Natl Acad Sci U S A [Internet]. 2014 Oct 20 [cited 2014 Oct 22];111(44):15798–803. Available from: http://www.ncbi.nlm.nih.gov/pubmed/25331901

27. Lucas C, Klein J, Sundaram M, Liu F, Wong P, Silva J, et al. Kinetics of antibody responses dictate COVID--19 outcome. medRxiv. 2020.

28. Dunning JW, Merson L, Rohde GGU, Gao Z, Semple MG, Tran D, et al. Open source clinical science for emerging infections. The Lancet Infectious Diseases. 2014. p. 8–9.

29. Amanat F, Stadlbauer D, Strohmeier S, Nguyen THO, Chromikova V, McMahon M, et al. A serological assay to detect SARS--CoV--2 seroconversion in humans. Nat Med. 2020;

